# Does Emergency Department point-of-care ultrasound in the evaluation of possible small bowel obstruction lead to meaningful improvements in patient-centric milestones?

**DOI:** 10.1101/2024.07.01.24309689

**Authors:** Yi-Ru Chen, Melva Morales Sierra, Rida Nasir, Naya Mahabir, Lisa Iyeke, Lindsay Jordan, Trupti Shah, Kevin Burke, Matthew Friedman, Daniel Dexeus, Athena Mihailos, Mark Richman, Joshua Guttman

**Affiliations:** Northwell Health Long Island Jewish Medical Center, Department of Emergency Medicine, 270-05 76th Ave., New Hyde Park, NY 11040; Associate Professor of Emergency Medicine, Emory University School of Medicine, Department of Emergency Medicine, 1273 Merry Lane NE Atlanta, GA 30329

## Abstract

**Introduction:** Point-of-care ultrasound (POCUS) has 90-95% sensitivity and specificity for small bowel obstruction (SBO) compared with computed tomography (CT). ED clinicians might reasonably use a positive POCUS to progress to patient-centric milestones (eg, nasogastric tube (NGT) placement, general surgery consult, and disposition). Awaiting CT performance and interpretation before moving to such milestones may delay care. Literature is limited concerning the effects of POCUS vs. CT alone on such patient-centric milestones for patients with SBO. This study compared time to patient-centric milestones (NGT, general surgery consult, and disposition) among ED patients suspected of having SBO who underwent POCUS vs. CT only in their SBO diagnostic process.

**Methods:** Data from 11,801 SBO patients seen among 14 EDs between 2017-2022 was queried. Patients were categorized into two groups according to diagnostic method (POCUS + CT vs. CT alone). Patients were included if they had a POCUS positive for SBO and an ED diagnosis of SBO; they were excluded from analysis of any specific/particular milestone (NGT, general surgery consult, or disposition) if they had that milestone prior to POCUS. Median time from ED arrival to each milestone was calculated for both groups (POCUS + CT vs. CT alone).

**Results:** Compared to CT-only patients, patients with POCUS plus CT had a non-statistically-significant longer wait time from ED arrival to NGT (414 vs. 390, p=0.7) and from ED arrival to general surgery consult (487.5 vs. 442 minutes, p = 0.07). They had statistically-significantly longer time to from ED arrival to disposition (475.5 vs. 377 minutes, p=0.009). Among cases in which POCUS was performed, 80% of the time the NGT was placed, 77% of the time the general surgery consult was performed, and 100% of time disposition was made only after CT result rather than after POCUS but before CT result.

**Conclusion:** Use of POCUS was not associated with earlier achievement of patient-centric milestones (NGT or general surgery consult) and was associated with longer time to disposition. This is most-likely because, despite POCUS suggesting SBO, clinicians waited for CT results prior to placing the NGT, consulting general surgery, and entering the disposition. Such results suggest that, despite POCUS’s high sensitivity and specificity, ED and/or general surgery clinicians rely on CT scan results to confirm SBO, delaying patient-centric milestones.

## Introduction

As an immediate, non-invasive, relatively-cheap, harmless, radiation-free procedure, Emergency Department (ED) bedside point-of-care ultrasound (POCUS) is frequently used to aid diagnosis and guide treatment decisions [1], including for small bowel obstructions (SBO). POCUS imaging is often accompanied with CT scanning, as CT scans are the radiographic “gold standard” for SBO, as well as numerous other conditions (eg, abdominal aortic aneurysm, renal stone) [2-3] that may be detected if SBO is not the patient’s actual condition. Patients undergoing CT may have a delay in care due to time to CT performance and interpretation, and receive radiation and, often, intravenous contrast exposure (which has been associated with anaphylaxis [4] and nephrotoxicity [5]). Radiation can be particularly-harmful to vulnerable populations (eg, children, fetuses). For these reasons, POCUS or radiology-performed ultrasound by healthcare clinicians is often preferred to other diagnostic procedures (eg, x -rays, CT) for diagnosis of certain conditions such as pediatric acute appendicitis [6].

However, POCUS use for the diagnosis of SBO has several potential disadvantages. Being operator- and interpreter-dependent, POCUS may lead to an increased risk of confirmation bias, misdiagnosis, and false assurance. Additionally, the quality of the examination may be impacted by a number of patient characteristics such as obesity, pregnancy, and presence of excessive intestinal gas [7]. This can lead to additional unnecessary testing (with associated risks and costs) due to false positive and negative findings.

Previous research has demonstrated POCUS is associated with earlier time to patient-centered care milestones/interventions for various conditions. Among patients in respiratory distress, POCUS decreased ED diagnostic time by two hours compared to radiography, CT scan, and echocardiography [8]. Another study that examined dyspnea using POCUS revealed an earlier time to diagnosis, with a decrease of 4.5 hours compared to other modes of imaging (eg, x-ray CT) [9]. An analysis of POCUS in patients with undifferentiated hypotension revealed an increase in the percent with a definitive diagnosis and significant changes in management plans (use of IV fluids, vasoactive agents, or blood products, consultation, major diagnostic imaging, and disposition) [10]. Lastly, a study of patients with pericardial effusions found a decreased time to accurate diagnosis, as well as a trend toward decreased hospital length-of-stay [11].

Recent studies have described and quantified the benefit of POCUS in changing management, as represented by the number needed to scan (NNS): the number of POCUS studies that need to be performed to result in one patient having: 1) a change in management, 2) improved procedural safety and accuracy, or 3) a diagnosis that would have been missed. Amini, et. al.’s systematic review found that, in affecting a course of management: 1) NNS = 4 to decide to incise and drain skin/soft tissue infections (ie, found an abscess that wasn’t clinically-obvious), 2) NNS = 2 to decide whether to perform arthrocentesis, 3) NNS = 4 to decide whether or not to give intravenous fluids for hypotension [12]. However, Amini’s investigation did not address small bowel obstructions (SBOs), where POCUS might aid in the decision to perform patient-centric milestones such as time to nasogastric tube (NGT) placement, general surgery consultation, or admission.

SBOs are increasingly-common in the United States, comprising an estimated 2-4% of ED abdominal pain visits [14] and 300,000 annual admissions [15] and accounting for around 12-16% of all surgical admissions [16].

POCUS has high sensitivity and specificity (∼90-95%) for identifying SBO [17]. However, only a small amount of literature exists regarding the beneficial effects of ED POCUS on patient-centric milestones (eg, time to surgical consultation) among patients with POCUS vs. those with CT. One study found ED POCUS positively impacts patients with SBO, reducing ED length-of-stay by two hours compared to patients diagnosed via a radiology-performed ultrasound [3]. Guttman et. al. concluded POCUS could eliminate the need for x-ray imaging [13]. Use of POCUS to exclude SBO could be particularly-useful in patients in whom the clinician has a low pretest probability for SBO.

This study investigated the time to patient-centric milestones (NGT, general surgery consult, and disposition) in ED patients with CT-demonstrated small bowel obstruction (SBO) who had vs. did not have POCUS prior to CT, to determine whether POCUS before CT led to changes in time to such milestones.

## Methods

Northwell Health is a 22-hospital health system largely operating in Long Island and New York City. Northwell’s EDs see approximately 850,000 visits/year. Patients presenting with possible SBO are triaged to an ED treatment area, where they are seen by a clinician. The clinician suspicious of an SBO typically orders a CT scan of the abdomen/pelvis. Depending on clinical suspicion and wait time for CT scan and general surgery consult, the clinician may either place an NGT themselves, or may wait for the CT result and general surgery before an NGT is placed. Clinicians comfortable with POCUS for SBO may perform a POCUS and then order the CT, after which they may either place an NGT themselves, or may wait for the CT result and general surgery.

Data from 14 Northwell EDs was queried from the Emergency Medicine Service Line covering April 2017 through December 2022. Patients were categorized into two groups according to diagnostic method (POCUS + CT vs. CT alone). Patients in the POCUS + CT group were included if they had a POCUS and CT positive for SBO and an ICD-10 ED diagnosis of SBO; patients in the CT-alone group were included if they had a CT positive for SBO and an ICD-10 ED diagnosis of SBO. We allowed for the possibility that POCUS might have been performed before the CT was performed, or after it was performed but before it was read by the Attending radiologist. However, we excluded patients whose POCUS was after CT was read. Patients were also excluded from analysis of any specific/particular milestone (NGT, general surgery consult, or disposition) if they had that milestone prior to POCUS. Median time from ED arrival to each milestone was calculated for both groups (POCUS + CT vs. CT alone). Among patients who underwent POCUS for SBO, we subtracted from the milestone timeframes the approximately 10 minutes it takes to organize, perform, and document POCUS.

This study was approved by the Northwell Health Institutional Review Board (IRB) (IRB #: 16-844).

## Results

Compared to CT-only patients, patients with POCUS plus CT had a non-statistically-significant longer wait time from ED arrival to NGT (414 vs. 390 minutes, p=0.7) and from ED arrival to general surgery consult (487.5 vs. 442 minutes, p = 0.07). They had statistically-significantly longer time to from ED arrival to disposition (475.5 vs. 377 minutes, p=0.009). Among cases in which POCUS was performed, 80% of the time the NGT was placed, 77% of the time the general surgery consult was performed, and 100% of time disposition was made only after CT result rather than after POCUS but before CT result. **(Table 1, Figure 1)**

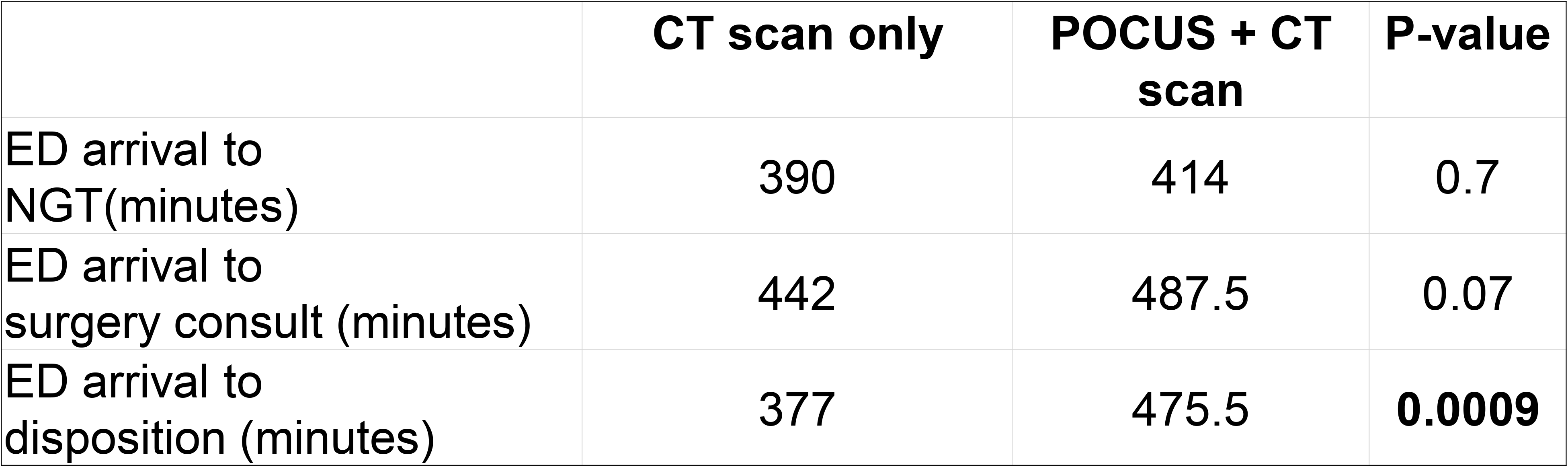

|  | CT scan only | POCUS + CT scan | P-value |
| --- | --- | --- | --- |
| ED arrival to NGT(minutes) | 390 | 414 | 0.7 |
| ED arrival to surgery consult (minutes) | 442 | 487.5 | 0.07 |
| ED arrival to disposition (minutes) | 377 | 475.5 | **0.0009** |

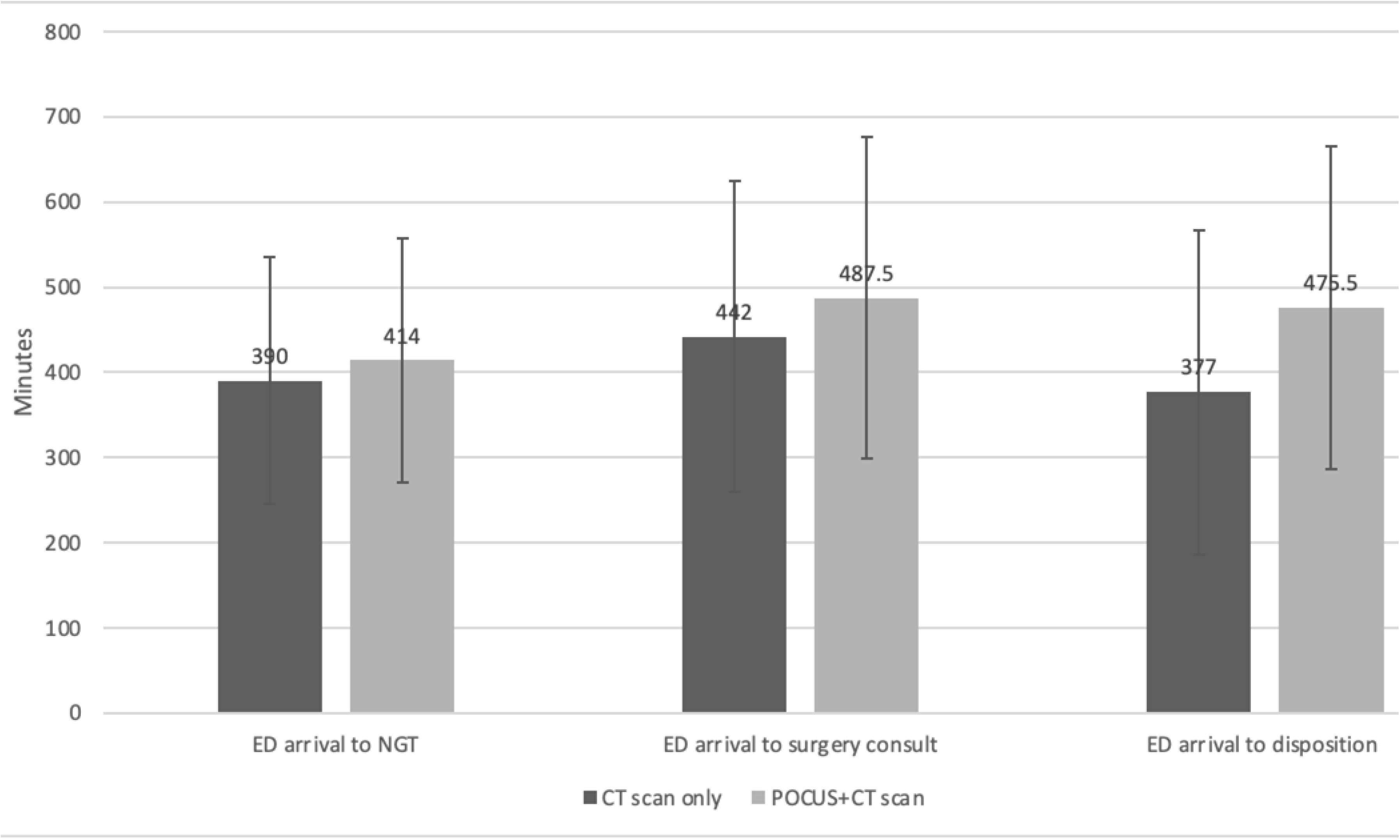

## Discussion

Use of POCUS was not associated with earlier achievement of patient-centric milestones (NGT or general surgery consult) and was associated with longer time to disposition. This is most-likely because, despite POCUS suggesting SBO, clinicians waited for CT results prior to placing the NGT, consulting general surgery, and entering the disposition. Such results suggest that, despite POCUS’s high sensitivity and specificity, ED and/or general surgery clinicians rely on CT scan results to confirm SBO, delaying patient-centric milestones.

In a previous study, we determined that, in a cohort of 106 patients who had POCUS to evaluate for SBO, the positive predictive value (PPV) of a positive POCUS was 73% (with a 23% prevalence of SBO among this cohort). With a 73% PPV, certain clinicians may be comfortable making clinical decisions to progress to NGT placement, general surgery consult, or disposition. However, others might not be, as our current study appears to demonstrate.

Although we did not perform chart review or query ED providers or general surgeons regarding their reliance on positive POCUS findings for SBO to guide next steps, our personal experience is that general surgeons often request CT following a positive POCUS, for many reasons, including localizing the site of obstruction; others have corroborated such observations [18].

### Limitations

This study has several limitations. First, this was not a randomized, controlled study. Patients may have undergone POCUS selectively (ie, selection bias), wherein the same clinician or patient characteristics which influenced who underwent POCUS also affected longer time to milestones. For example, older patients might have been more-likely to have both POCUS and longer ED work-ups because of complexity associated with aging. Alternatively, less-sick patients may have had POCUS as a screening examination and, when a “surprise” SBO was found on POCUS, also to have less-rapid patient-centric milestones, as they were less sick (ie, did not have as timely a need for NGT). Second, we performed a univariate (not multivariate) analysis; we were, therefore, unable to isolate which variables (eg, acuity) may have been associated with obtaining a POCUS or having a longer time to milestones. In addition, this study was conducted in one health system (Northwell). Finally, we did not conduct chart review, so we are unable to tell exactly why the clinician did not act on positive POCUS results for SBO.

## Conclusion

Despite having excellent sensitivity and specificity, and very good positive predictive value for SBO, a positive POCUS for SBO is not associated with decreased time to patient-centric milestones (NGT, general surgery consult, or admission). This suggests ED and surgery providers likely do not act immediately upon positive POCUS SBO results and, instead, wait for confirmatory CT scans.

## Data Availability

All data produced in the present work are contained in the manuscript

